# Repurposing azacitidine and carboplatin to prime for anti-PDL1 re-challenge of immunotherapy-resistant melanoma

**DOI:** 10.1101/2022.02.03.22270286

**Authors:** Andre van der Westhuizen, Megan Lyle, Moira C. Graves, Xiaoqiang Zhu, Jason W. H. Wong, Kerrie Cornall, Shu Ren, Leanna Pugliese, Richard Levy, Adeeb Majid, Ricardo E. Vilain, Nikola A. Bowden

## Abstract

Drug repurposing offers the opportunity for approved chemotherapy agents to be used to re-establish sensitivity to immune checkpoint blockade (ICB) therapy. Here we investigated the clinical and translational aspects of an early phase II study of azacitidine and carboplatin priming for anti-PDL1 immunotherapy (Avelumab) in patients with advanced ICB-resistant melanoma. 20 participants with ICB resistant metastatic melanoma received 2 × 4-week cycles of azacitidine and carboplatin followed by ICB re-challenge with anti-PD-L1 avelumab. The overall response rate (ORR) determined after 2 × 4-week cycles of azacitidine and carboplatin priming was 10% (2/20) with 2 partial responses (PR). The ORR determined after priming followed by 6 cycles of avelumab (week 22) was 10%, with 2/20 participants achieving iPR. The clinical benefit rate (CBR) for azacitidine and carboplatin priming was 65% (13/20) and after priming followed by 6 cycles of avelumab CBR was 35% (n = 7/20). The median PFS was 18.0 weeks (95% CI: 14.87 – 21.13 weeks) and the median OS was 47.86 weeks (95% CI: 9.67 – 86.06 weeks). Translational correlation analysis of tumour biopsies at baseline, after priming and after 6 cycles of avelmuab confirmed HLA-A generally increased after priming with azacitidine and carboplatin, particularly if it was absent at the start of treatment. Average methylation of CpGs across the HLA-A locus showed a consistent decrease in methylation after priming and T-cells, in particular CD8+, showed the greatest increase in infiltration. Priming with azacitidine and carboplatin can induce disease stabilization and re-sensitisation to ICB for metastatic melanoma.

**One Sentence Summary:** Sequential azacitidine and carboplatin stabilises disease burden and re-establishes sensitivity to checkpoint immune blockade immunotherapy.

## INTRODUCTION

Despite significant progress in the treatment of metastatic melanoma with molecularly targeted and immune checkpoint blockade (ICB) therapy in recent years, there remains limited effective treatment once resistance occurs. The mechanisms of resistance to ICB are being extensively explored, and recently the down-regulation of human leukocyte antigen A (HLA-A) expression and other key immune pathways have been identified as a key component in the development of ICB resistance (*1, 2*).

Drug repurposing offers the opportunity to determine if more affordable, off-patent and FDA/TGA approved chemotherapy agents can be used to increase immune response pathways and re-establish sensitivity to ICB immunotherapy. The mechanism of ‘priming’ aims to re-establish immune response in ICB resistant patients. Epigenetic and DNA-damaging chemotherapy have long been known to induce immune-related responses in solid tumours, but used together they have the potential to synergistically prime to re-instate the response to ICB.

DNA methylation plays an important role in cancer and commonly alters the expression of tumour suppressor genes in the absence of somatic mutations (*3*). In cutaneous melanoma, global hypomethylation increases DNA instability and local hypermethylation of promoter CpG islands can silence the expression of tumour suppressor genes (*4*). These changes lead to alterations in pathways, including cell cycle regulation, cell signalling, transcription, DNA repair and apoptosis (*5*).

DNA methylation provides an appealing target for drug repurposing to overcome ICB resistance due to the established and safe use of DNA hypomethylating agents such as 5-aza-5’deoxycytidine (decitabine) and azacitidine. DNA hypomethylating agents are cytosine analogues that bind to DNA methyltransferase 1 (DNMT1) and inhibit the function of maintaining DNA methylation during cell division, leading to passive demethylation with replication (*6*). The inhibition of methylation restores the expression of silenced genes (*7*). Azacitidine is used primarily in the treatment of haematological malignancies, including myelodysplastic syndrome and chronic myeloid leukaemia but has since become a useful mechanism for studying the effect of methylation on gene expression (*8*). At high doses both decitabine and azacitidine can be cytotoxic, inducing DNA damage from DNTM1 persistently bound to DNA, but at low doses, can effectively demethylate without causing cell death (*9, 10*).

Pre-clinical studies and clinical trials have investigated the therapeutic potential of DNA hypomethylating agents as monotherapy, and in combination, in several solid cancer types with positive results. Of the 16 most commonly used chemotherapy agents, DNA-damaging cisplatin and carboplatin show the best synergy with decitabine in vitro (*11*). This synergy affects transcriptional activation of silenced genes, with the addition of carboplatin increasing gene expression from a 10-fold increase to a 32-fold increase (*11*).

To date, the combination of hypomethylating and DNA-damaging agents has been reported in pre-clinical studies (*12*) but has not been assessed by clinical trials in melanoma. The combination has however, shown promising effects in ovarian cancer, with high response rate and progression-free survival in platinum-resistant patients (*13*). Similar results have been found in head and neck squamous cell carcinoma(*14*), neuroblastoma (*15*), renal carcinoma (*16*) and non-small cell lung cancer (*17*).

Here we investigated the clinical and translational aspects of the ‘priming’ combination of hypomethylating and DNA-damaging agents in an early phase II study of azacitidine and carboplatin priming for anti-PDL1 immunotherapy (Avelumab) in patients with advanced melanoma who were resistant to ICB (PRIME002: ACTRN12618000053224).

## RESULTS

### Cohort characteristics

Participants with metastatic melanoma (n=20) were recruited between March 2018 and December 2020. All participants had confirmed disease progression whilst treated with single agent anti-PD1 ICB (n=12) or combination anti-CTLA4 and anti-PD1 ICB (n=8). 8 participants had also received targeted BRAF/MEK inhibitors prior to ICB. The average number of lines of previous treatment was 2, with a range of 1-8 lines.

The average age was 63.9 ± 14.4 years and the cohort consisted of 4 females and 16 males. ECOG scores were all 0 or 1, with the exception of ECOG score 2 for participant 017. A summary of the cohort characteristics is presented Table 1.

**Table 1.** Cohort characteristics.

| **Patient** | **Age Category** | **Gender** | **BRAF** | **# Lines** | **ECOG** |
| --- | --- | --- | --- | --- | --- |
| 001 | <65 | F | WT | 1 | 0 |
| 002 | >65 | M | WT | 1 | 1 |
| 003 | <65 | M | V600 | 2 | 0 |
| 004 | >65 | M | WT | 1 | 0 |
| 005 | >65 | M | WT | 1 | 0 |
| 006 | <65 | F | WT | 2# | 0 |
| 007 | >65 | M | WT | 2# | 0 |
| 008 | >65 | M | WT | 2# | 0 |
| 009 | <65 | M | V600 | 5# | 1 |
| 010 | >65 | M | WT | 1 | 1 |
| 011 | >65 | M | WT | 2 | 1 |
| 012 | <65 | F | V600 | 4# | 0 |
| 013 | <65 | M | V600 | 6 | 1 |
| 014 | <65 | M | V600 | 4 | 0 |
| 015 | >65 | M | WT | 3 | 1 |
| 016 | >65 | M | WT | 1 | 1 |
| 017 | <65 | M | V600 | 8# | 2 |
| 018 | >65 | M | V600 | 2 | 0 |
| 019 | <65 | F | V600 | 2# | 1 |
| 020 | <65 | M | WT | 3# | 1 |

#anti-CTLA4 and anti-PD1 checkpoint inhibitor failure

### Objective Response rate (ORR)

The primary outcome measure of the study was the objective response rate (ORR) after 2 cycles of priming according to RECIST 1.1 (*18, 19*) and then after 6 cycles of immunotherapy (Avelumab) rechallenge according to iRECIST (*20*). The ORR of epigenetic and DNA-damaging “priming’ treatment was determined after 2 × 4-week cycles of azacitidine and carboplatin priming (week 9) was 10% (2/20) with 2 partial responses (PR). No complete responses (CR) occurred.

The ORR of epigenetic and DNA-damaging priming followed by immunotherapy re-challenge determined after 2 × 4-week cycles of azacitidine and carboplatin priming followed by 6 cycles of anti-PD-L1 immunotherapy (Avelumab) (week 22) was 10%, with 2/20 participants achieving iPR. Interestingly, the 2 participants with PR after azacitidine and carboplatin priming did not maintain the PR during immunotherapy rechallenge. The 2 participants with iPR after 6 cycles of Avelumab had SD after priming with responses close to PR at -28.1% and -29.7%.

### Clinical Benefit Rate (CBR)

The secondary outcome measure of the study was clinical benefit rate (CBR) determined as the percentage of participants with CR, PR or stable disease (SD) after 2 × 4-week cycles of azacitidine and carboplatin priming (week 9) and then after 6 cycles of immunotherapy (Avelumab) rechallenge. In addition to the 10% (2/20) PR, 55% (11/20) maintained SD resulting in a clinical benefit rate (CBR) for azacitidine and carboplatin priming of 65% (13/20).

The CBR of epigenetic and DNA-damaging priming followed by immunotherapy re-challenge was determined after 2 × 4-week cycles of azacitidine and carboplatin priming followed by 6 cycles of anti-PD-L1 immunotherapy (Avelumab) according to iRECIST. In addition to the 10% iPR (2/20), 25% (5/20) maintained SD resulting in CBR of 35% (n = 7/20).

### Tumour response to epigenetic and DNA-damage priming for immunotherapy re-challenge

Disease stabilization or partial response was achieved for RECIST 1.1 measurable target lesions for 94.4% (17/18) patients that completed 2 × 4-week cycles of epigenetic and carboplatin priming (Figure 1A). 2 patients rapidly progressed and did not complete the 2 cycles of azacitidine and carboplatin priming. 3 patients with stable target lesions had confirmed PD according to RECIST 1.1 criteria due to unequivocal progression of non-target lesions (n = 2) or a new lesion (n = 1).

**Figure 1:**
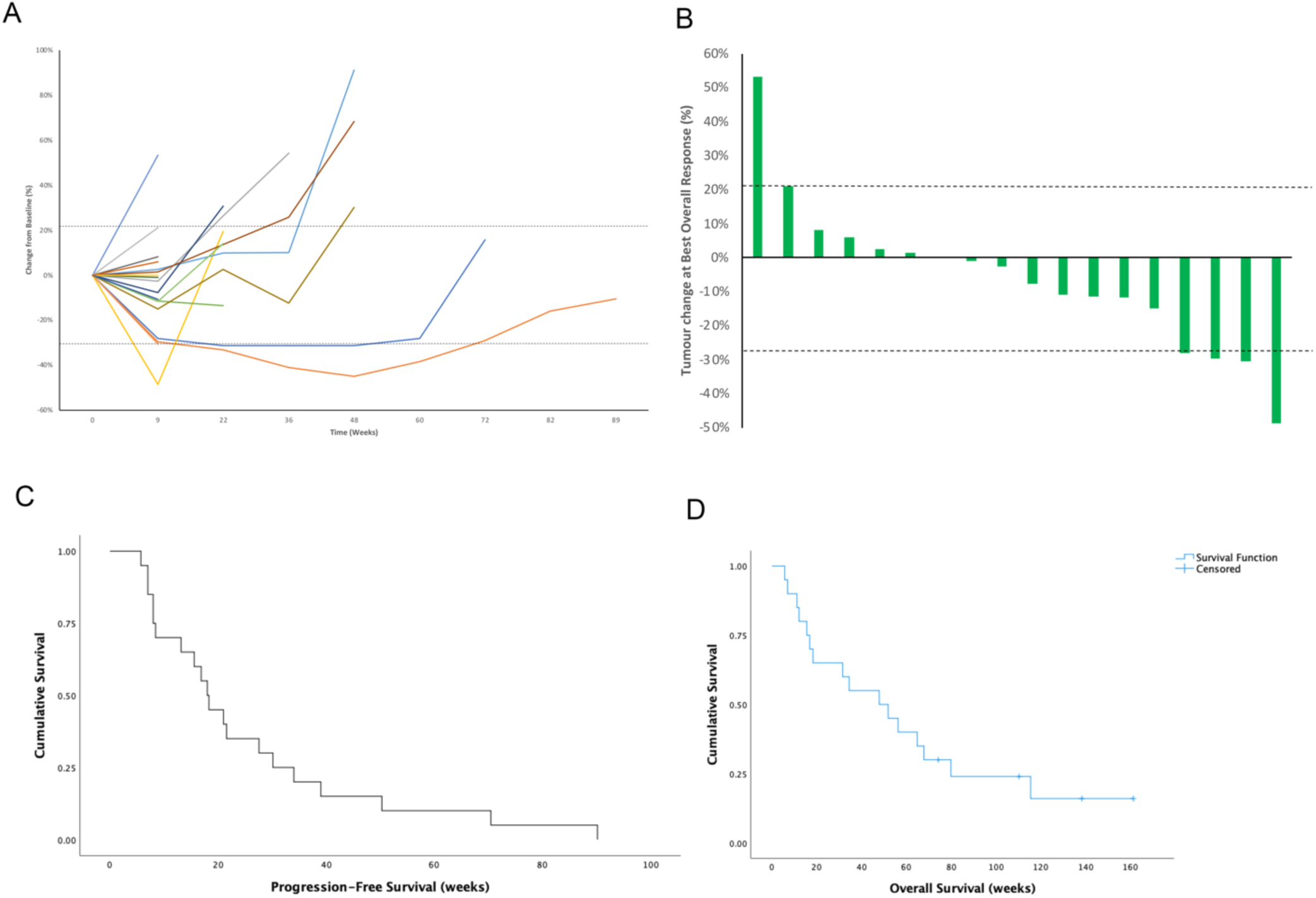
Tumour response, progression-free survival and overall surivival. A) Spider plot of tumor burden change of target lesions in reference to the baseline tumur burden in 18 patients with measurable tumor burden; B) Waterfall plot of the tumor burden change of target lesions at best response (%) in reference to the baseline tumor burden in 18 patients with measurable tumor burden. Dashed lines at +20% and − 30% represent the threshold used for progression (PD) and partial response (PR); C) Progression-Free Survival (PFS) in weeks. The median PFS from start of treatment was 18.0 weeks (95% CI: 14.87 – 21.13 weeks); D) Overall Survival (OS) in weeks. The median OS was 47.86 weeks (95% CI: 9.67 – 86.06 weeks)

Disease stabilization or partial response was achieved for iRECIST measurable target lesions for 77.8% (7/9) patients that completed 2 × 4-week cycles of epigenetic and carboplatin priming followed by 6 cycles of Avelumab (Figure 1A). 55% (11/20) patients had confirmed PD before week 22.

### Progression Free Survival (PFS) and Overall Survival (OS)

The median PFS from the start of cycle 1 of azacitidine and carboplatin was 18.0 weeks (95% CI: 14.87 – 21.13 weeks) (Figure 1C). The proportion of PFS at week 9 after 2 cycles of priming was 65% (13/20) of participants, which decreased to 35% (7/20) by week 22 (Table 2).

**Table 2.**
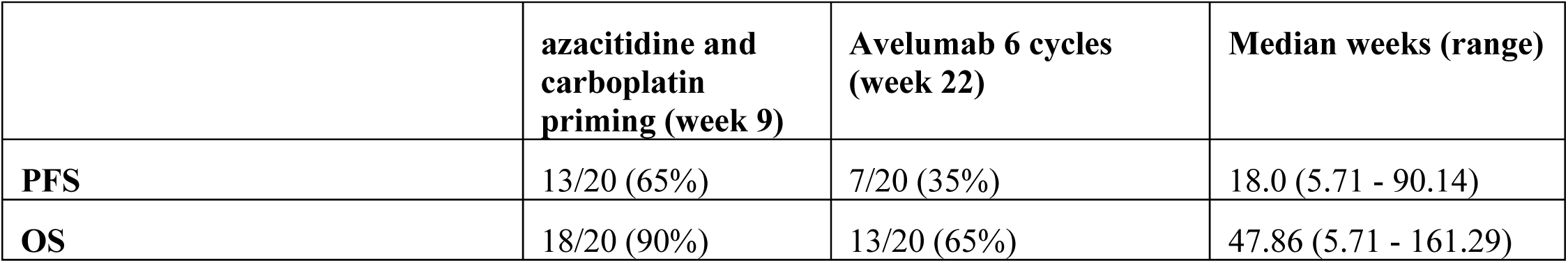
Progression-free survival (PFS) and overall survival (OS)

|  | **azacitidine and carboplatin priming (week 9)** | **Avelumab 6 cycles (week 22)** | **Median weeks (range)** |
| --- | --- | --- | --- |
| **PFS** | 13/20 (65%) | 7/20 (35%) | 18.0 (5.71 - 90.14) |
| **OS** | 18/20 (90%) | 13/20 (65%) | 47.86 (5.71 - 161.29) |

The median OS from the start of cycle 1 was 47.86 weeks (95% CI: 9.67 – 86.06 weeks) (Figure 1D). 90% (18/20) of participants survived beyond week 9 and 65% (13/20) beyond week 22 (Table 2).

### Assessment of safety

No grade 4 or persistent grade 3 treatment-related adverse events (TRAEs) occurred as result of the azacitidine and carboplatin priming or re-challenge with Avelumab. All reported grade 1-3 TRAEs are summarized in Table 3.

**Table 3.** Treatment related adverse events (TRAEs)

| **CTCAE Grade** | **Treatment Related Adverse Event (TRAE)** | **Count** | **Patients (n)** | **Patients (%)** |
| --- | --- | --- | --- | --- |
| Grade 4 |  | 0 | 0 | 0 |
| Grade 3 | Neutropenia<sup>#</sup> | 2 | 2 | 10% |
|  | Infusion Reaction<sup>#</sup> | 1 | 1 | 5% |
|  | Pneumontis<sup>#</sup> | 1 | 1 | 5% |
|  | Fever<sup>#</sup> | 1 | 1 | 5% |
| Grade 2 | Mucositis | 1 | 1 | 5% |
|  | Infusion Reaction | 1 | 1 | 5% |
|  | Rash | 1 | 1 | 5% |
|  | Neutropenia | 1 | 1 | 5% |
|  | Pneumonitis | 1 | 1 | 5% |
|  | Diarrhea/Colitis | 1 | 1 | 5% |
|  | Fevers | 1 | 1 | 5% |
|  | urinary tract infection | 1 | 1 | 5% |
|  | Fatigue | 1 | 1 | 5% |
| Grade 1 | Neutropenia | 4 | 4 | 20% |
|  | Fatigue | 4 | 4 | 20% |
|  | Nausea | 4 | 4 | 20% |
|  | Lethargy | 3 | 2 | 15% |
|  | Dysgeusia | 4 | 3 | 15% |
|  | constipation | 2 | 2 | 10% |
|  | Headaches | 2 | 2 | 10% |
|  | Hepatitis | 2 | 2 | 10% |
|  | Decreased Lymphocytes/white cell count | 2 | 1 | 5% |
|  | anaemia | 1 | 1 | 5% |
|  | Hypomagnesemia | 1 | 1 | 5% |
|  | Joint symptoms | 1 | 1 | 5% |
|  | loose stools | 1 | 1 | 5% |
|  | Metallic taste- dysgeasia | 1 | 1 | 5% |
|  | Platelet Count Decreased | 1 | 1 | 5% |
|  | vomiting | 1 | 1 | 5% |
|  | weight loss | 1 | 1 | 5% |
|  | Diarrhea/ Colitis | 1 | 1 | 5% |
|  | Rash | 1 | 1 | 5% |

<sup>#</sup>duration <7 days

Whole blood counts were collected at baseline, week 9 (after 2 cycles of azacitidine and carboplatin) and week 22 (after 6 cycles of Avelumab). There was no significant difference in whole blood cell counts at week 9 or week 22 compared to baseline (supplementary figure 1).

### Translational aspects of priming mechanism and immunotherapy rechallenge: Expression and localization of HLA-A after epigenetic and carboplatin priming and immunotherapy re-challenge

Biopsies were limited to 4 participants as tissue collection was restricted to accessible nodal or subcutaneous lesions and where the lowest risk of morbidity was expected in doing the biopsy. Despite the small sample size, HLA-A expression and lymphocyte infiltrate was investigated at baseline, after priming (week 9) and after ICB re-challenge (week 22). HLA-A generally increased after priming with azacitidine and carboplatin and re-challenge with Avelumab, particularly if it was absent at the start of treatment (Figure 2a, b, c).

**Figure 2.**
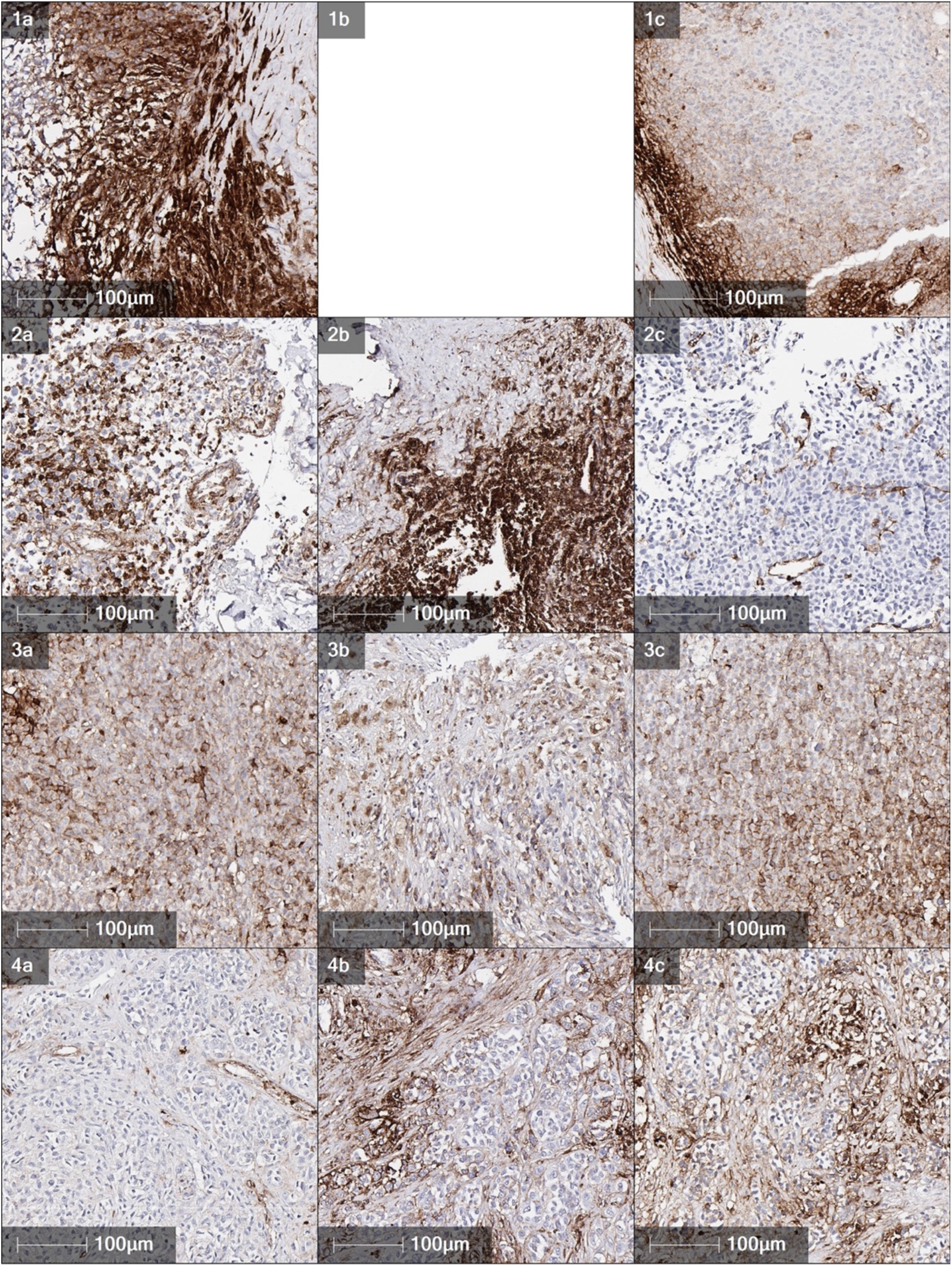
HLA-A immunohistochemistry in baseline (1a, 2a, 3a, 4a), week 9 (1b, 2b, 3b, 4b) and week 22 (1c, 2c, 3c, 4c) biopsies. HLA-A generally increased after priming with azacitidine and carboplatin and re-challenge with Avelumab, particularly if it was absent at the start of treatment

### Altered DNA methylation and gene expression of antigen presentation and immune signalling after epigenetic and carboplatin priming and immunotherapy re-challenge

Epigentic (WGBS) and transcriptomic (RNA-seq) profiling were performed for three patients (Pt 6, Pt 8 and Pt 10). These patients all had SD at week 9, and at week 22, biopsies were measured as SD by iRECIST, but Pt 8 and 10 had 1 new lesion at week 22 and were iCPD. Frozen biopsies were available for all time points for the three patients except at week 22 for Pt 10. We first assessed the impact of genome-wide DNA methylation changes induced by azacitidine treatment. CpGs were stratified in relation to CpG islands. As expected, most CpG islands are unmethylated, with shores partially methylated. CpG island shelves and non-island CpGs were mostly methylated (Figure 3a). Upon treatment, at week 9, there was a modest change in methylation level across all regions, with no noticeable trend for Pt 6 and Pt 10 across week 9 and week 22, while for Pt 8, there was a general decrease at week 9 and increase at week 22. Comparing unmethylated (< 30%, Fig 3b) and methylated (>30%, Fig 3c) CpG islands, it was found that decrease in methylation level at week 9 was consistent across all patients, suggesting that azacitidine has the greatest effect on DNA methylation at hypermethylated CpG islands.

**Figure 3.**
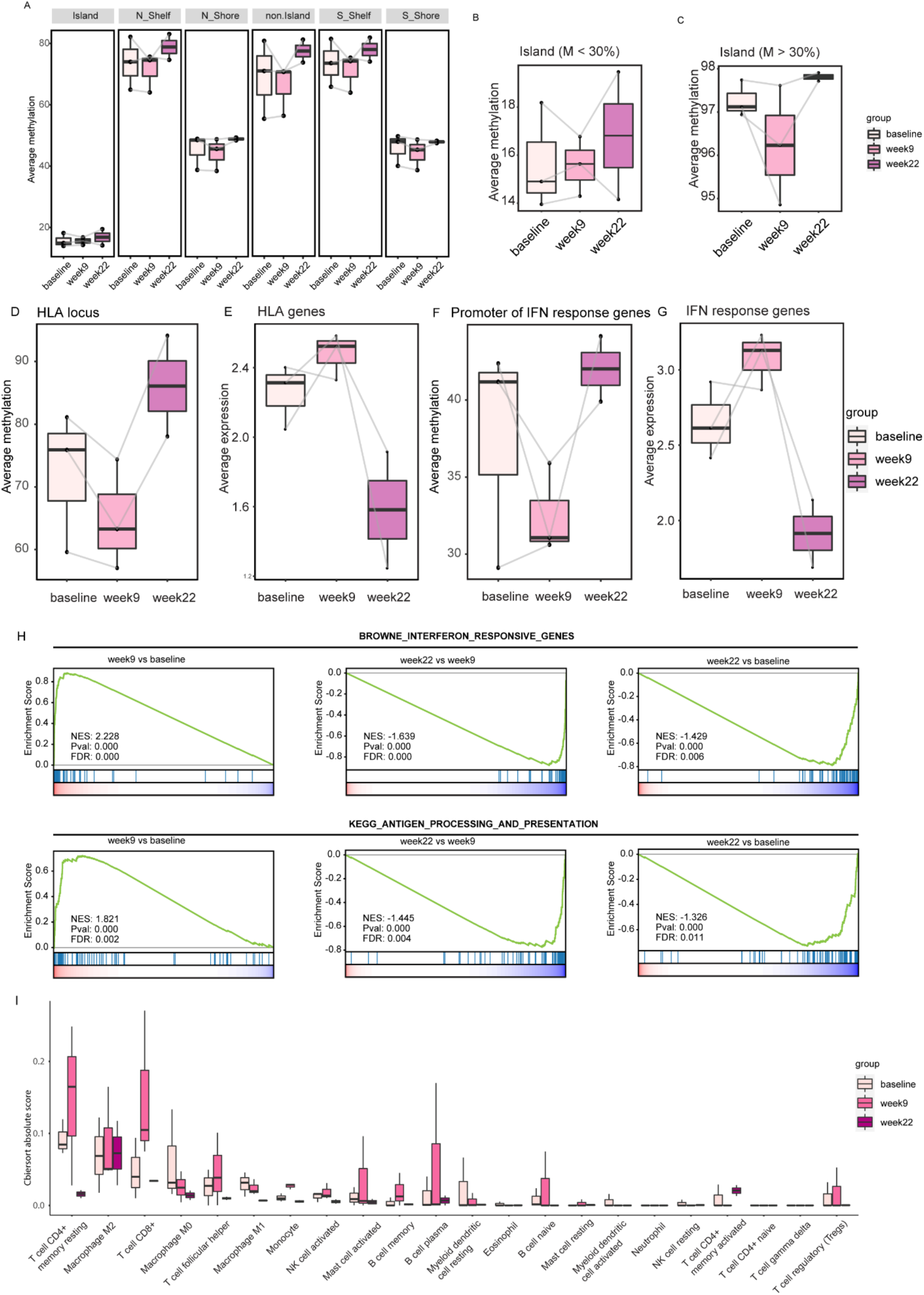
DNA methylation and gene expression analysis of patient samples at baseline, week 9 and week 22. Average methylation across different CpG loci in relation to CpG islands (A). Average methylation of unmethylated (B) and methylated (C) CpG islands. Average CpG methylation (D) and gene expression (E) change across HLA-A locus. Average CpG methylation change at interferon (IFN) response gene promoters (F) and gene expression change of corresponding IFN response genes (G). Geneset enrichment analysis of IFN response and antigen processing and presentation pathway for time point comparisons (H). CIBERSORT scores of immune cell infiltration at the different time points (I)

As immunohistochemistry had shown an increase in HLA-A staining following priming (week 9) (Fig 2), we sought to determine if this was related to DNA demethylation caused by azacitidine treatment. Using the WGBS data, we quantified the average methylation of CpGs across the HLA-A locus, which indeed showed a consistent decrease in methylation at week 9 in all three patients (Fig 3d). This demethylation was mirrored by an increase in the expression of HLA-A genes at week 9 (Fig 3e). To further assess the impact of azacitidine priming on immune response, we also assessed the impact of DNA demethylation on interferon (IFN) response genes. Two patients (Pt 6 and Pt 8) showed substantial average demethylation of the promoters of IFN response genes, while there was a slight increase in Pt 10 (Fig 3f). The impact of the change in DNA methylation was again reflected in the average expression of these genes (Fig 3g). More generally, using the gene expression data for geneset enrichment analysis (GSEA), we found a significant increase in the expression of IFN response and antigen processing and presentation pathways (Fig 3h) when comparing week 9 with baseline. Interestingly, this trend is reversed by week 22, which may reflect a change in the composition of both the tumour and its microenvironment.

Recent studies have shown that one of the mechanisms of action of azacitidine treatment is to induce the expression of transposable elements (TE) (*21*). Using the REdiscoverTE pipeline (*22*), we quantified changes in TE across the three time points. Interestingly, unlike IFN and antigen presentation, there was a sustained increase in the expression of most TE subfamilies even at week 22. However, this did not corroborate with average methylation change at these TE loci, which suggested that TE over-expression may be restricted to limited loci, making it difficult to detect by average methylation levels (Supplementary Fig 2).

Finally, we used CIBERSORT (*23*) to determine the level of infiltration of different immune cells in these tumours. At baseline, T-cells and macrophages had the highest CIBERSORT scores suggesting that they are the most dominant infiltrating immune cells in these tumours. Importantly, at week 9, it was found that T-cells, in particular CD8+, showed the greatest increase in infiltration. CD8+ T cells can indicate a “hot” tumour microenvironment and is a predictor of immunotherapy response (*23*). This is consistent with our hypothesis that azacitidine and carboplatin prime the tumour microenvironment for response to immune checkpoint blockade therapy.

### T-cell markers in peripheral blood after epigenetic and carboplatin priming and immunotherapy re-challenge

T-cell exhaustion proliferation and migration markers were quantified from peripheral blood using flow cytometry at baseline, week 9 after 2 cycles of azacitidine and carboplatin and week 22 after 6 cycles of Avelumab. There was no significant difference in the markers at week 9 or week 22 compared to baseline. There was an overall trend of a small increase after priming at week 9 which requires confirmation in a larger cohort (Figure 4).

**Figure 4.**
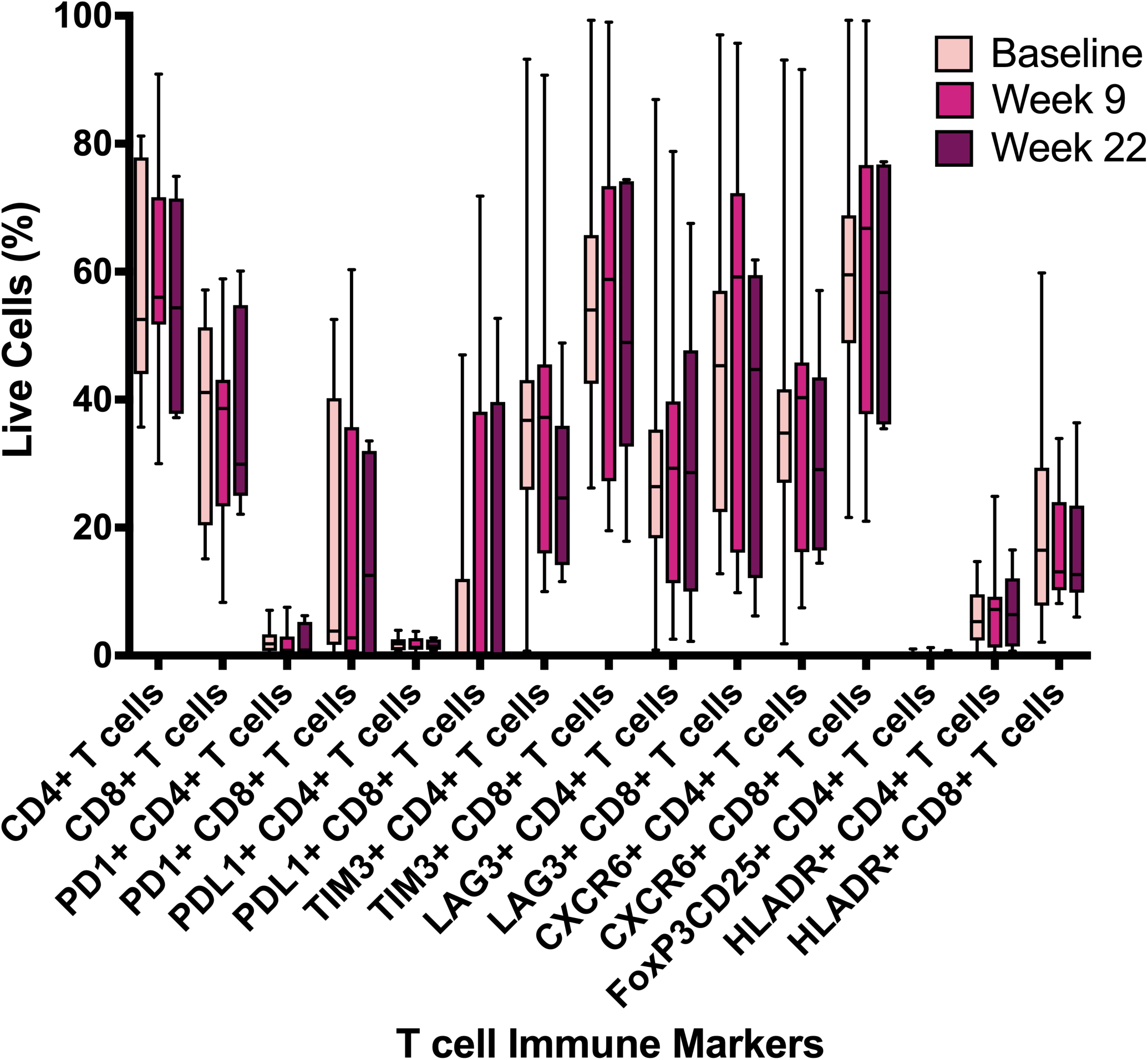
T-cell exhaustion proliferation and migration markers. Flow cytometry was used to quantify markers from peripheral blood at baseline, week 9 after 2 cycles of azacitidine and carboplatin and week 22 after 6 cycles of Avelumab.

## DISCUSSION

The clinical and translational data in this early phase II study suggests that the mechanism of priming with azactidine and carboplatin changes the tumour dynamics by enhancing the pathways related to antigen expression resulting in stabilisation of disease burden and response to ICB re-challenge. The regime was well tolerated as indicated by the low incidence and grade of toxicities (TRAE table reference) suggesting that priming does not need to be at maximum tolerated dose to be beneficial, especially in a late stage heavily pre-treated population with increasing disease burden and often poor performance status.

This study design consisted of 2 cycles of priming with azacitidine and carboplatin which resulted in stable disease for 65% (13/20) of patients and partial response in a further 10% of patients (2/20). The data suggests that more cycles of priming in future studies may need to be considered to improve clinical responses, antigen presentation and expression of immune related transcripts.

HLA-A encodes for an antigen-presenting major histocompatibility complex class I (MHCI) molecule. HLA-A complexes with beta 2 microglobulin (B2M) to present viral and tumor-derived peptides on antigen-presenting cells for recognition by alpha-beta T cell receptor (TCR) on CD8-positive T cells, guiding antigen-specific T cell immune response to eliminate cells. Increasing the expression of HLA-A may therefore overcome ICB resistance and allow previously resistant patients to be re-challenged with ICIs. The clinical problem we face is how to increase HLA-A expression in ICB resistant melanoma.

The highest level of antigen pathway expression occurred during or shortly after priming but returned to the same or lower levels by 6 cycles of Avelumab. The increase in antigen pathway expression may need to be captured by combining some or all of the priming cycles with ICB in future studies. The data also suggest that there is loss of expression of the antigen pathway over time which indicates that re-priming at 4-6 months intervals may be beneficial to maintain sensitivity to ICB.

The limited responses seen to single agent ICB re-challenge in this study suggest that increasing the number of priming cycles and overlapping with commencement of ICB may be beneficial. Re-challenge with combination anti-PD1/PDL1 and anti-CTLA4 may also be beneficial to improve responses in future studies, especially considering the impact of priming on HLA expression in this study and the mechanism of action of anti-CTLA4 ICB (*24*)

The data also suggests that other platinum resistant tumour groups may benefit from the priming mechanism to establish or re-establish ICB sensitivity. A recent study in high-grade serous ovarian cancer also investigated the combination of the DNA methyltransferase inhibitor guadecitabine and concluded that inhibiting DNA methylation altered gene expression related to DNA repair and immune activation and re-sensitised to carboplatin. There is high potential to further exploit the response to sensitise ovarian cancer to ICB(*25, 26*).

In conclusion, priming with azacitidine and carboplatin can induce disease stabilisation and re-sensitisation to ICB for metastatic melanoma. Future larger prospective clinical and translational studies are needed to further elucidate the role and mechanism of priming as a way to improve or re-establish responses to ICB.

## MATERIALS AND METHOD

### Study Design

#### Sample size calculation

Power calculation was used to determine the study was 80% powered (β=0.2) to detect an ORR of 20% (α = 0.05) for a sample size of 17. To ensure greater than 80% power was achieved the study was closed with a sample size of 20.

#### Research Subjects

This study was approved by the Northern Sydney Local Health District HREC, reference number HREC/17/HAWKE/55’ Australian clinical trial registry number: ACTRN12618000053224; registered 16th January 2018. 20 patients with unresectable or metastatic melanoma with primary resistance to immune checkpoint blockade immunotherapy were recruited to assess ORR, CBR and adverse events.

#### Research Objectives

Primary Objective: Quantify overall response rate (ORR) after 2 cycles of priming according to RECIST 1.1 and 6 cycles of immunotherapy (Avelumab) according to iRECIST criteria(*20*). Secondary Objectives: Quantify complete response (CR), partial response (PR), stable disease (SD) and clinical benefit rate (CBR) after administration of 2 cycles of azacitidine and carboplatin/28 day cycle using RECIST 1.1 criteria; Quantify CR, PR, SD and CBR after administration of 2 cycles of azacitidine and carboplatin/28 day cycle using RECIST 1.1 and 6 cycles of immunotherapy (Avelumab) according to iRECIST criteria; Determine genome-wide DNA methylation and transciptome changes after administration of 2 cycles of azacitidine and carboplatin and 6 cycles of immunotherapy (Avelumab); Quantify immune-response markers (PDL-1, PD-1, CD4/CD8, and CD68) in blood before treatment, after 2 priming cycles (azacitidine and carboplatin) and after 6 immunotherapy (Avelumab) cycles; Calculate progression-free survival (PFS) and overall survival (OS) at each RECIST data collection point (weeks -1, 9 and 22) and every 6 months until study completion.

#### Interventional early phase II study design

Patients were treated with an epigenetic and platinum chemotherapy priming program of 2 × 4 week cycles (Figure 5).

**Figure 5.**
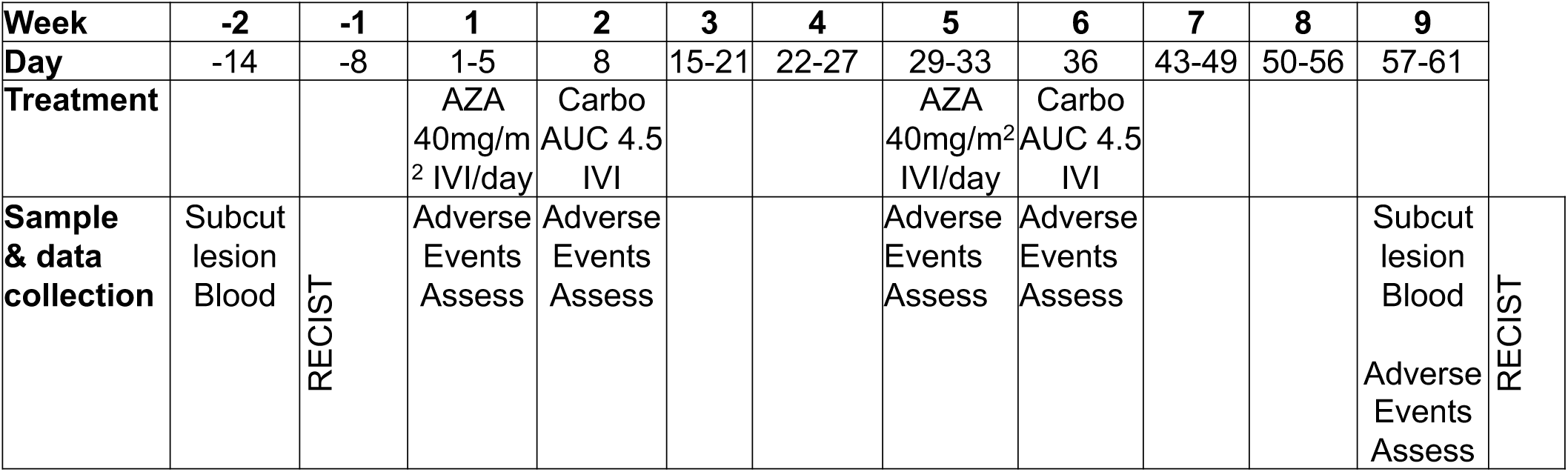
Epigenetic and platinum chemotherapy priming treatment schema.

Followed by an immunotherapy maintenance program until disease progression, death or withdrawal from study (Figure 6).

**Figure 6.**
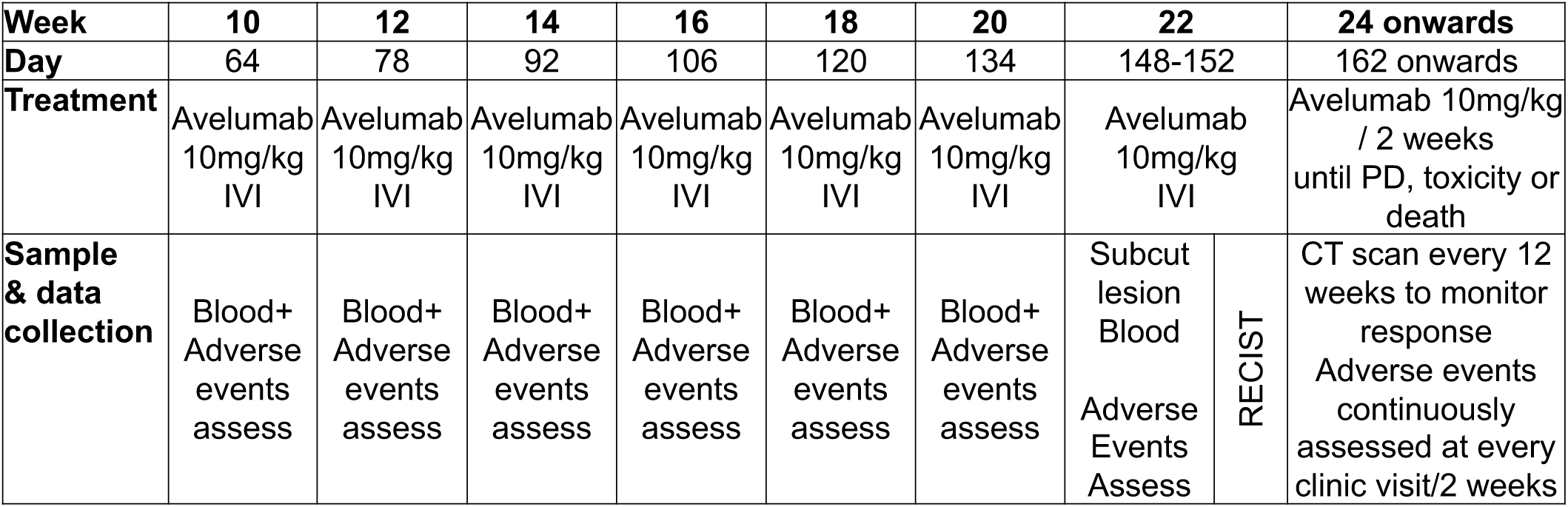
Immunotherapy treatment schema.

#### Primary outcome measures

Objective response rate (ORR) after 2 cycles of priming (1 cycle= azacitidine x 5 days followed by Carboplatin on Day 8/28 day cycle) according to RECIST 1.1 and 6 cycles (1 cycle = 1 dose of Avelumab/14 day cycle) of immunotherapy according to iRECIST criteria.

#### Secondary outcome measures

Complete response (CR), partial response (PR), stable disease (SD) and clinical benefit rate (CBR) after 2 cycles of priming (1 cycle= azacitidine x 5 days followed by Carboplatin on Day 8/28 day cycle) according to RECIST 1.1 and 6 cycles (1 cycle = 1 dose of Avelumab/14 day cycle) of immunotherapy according to iRECIST criteria.

### Translational experimental design

#### Immune cell profiles

Whole blood was collected at baseline, week 9 (after 2 cycles of azacitidine and carboplatin) and wee 22 (after 6 cycles of avelumab). Whole blood was separated into peripheral blood mononuclear cells (PBMCs) and plasma and stored at -80°C.

PBMCs were thawed rapidly, washed in RPMI media, rested for one hour at 37 ° C and washed twice more in RPMI. The cells were resuspended at a concentration of 1-10×10^6^ cells/mL, and 1uL of BD Horizon Fixable Viability stain 575V was added and incubated at room temperature for 15 minutes. Samples were washed twice with BD Pharmingen Stain Buffer. The following targets were co-stained: Hu CD3 BUV737 UCHT1, Hu CD4 BUV496 SK3, Hu CD8 APC-H7 SK1, Hu CD279(PD-1) BB515 EH12.1, Hu TIM-3(CD366) Alexa 647 7D3, Hu LAG-3(CD223) APC-R700 T47-530 was added to each sample and incubated for 30 minutes at 2-8 °C. The samples were washed twice with stain buffer. After final wash, 350uL of stain buffer was added to each sample and data acquired on a Fortessa X20 (BD Biosciences). Each sample was analysed in duplicate, non-consecutively.

Gated T-cells (CD3^+^, live, single cell, lymphocytes) were gated as CD4^+^ or CD8^+^ and further divided by presence of PD-1, LAG3 or TIM3. Co-expression of the 3 exhaustion markers was assessed but no further analysis was conducted due to very low percentage of cells positive for 2 or more exhaustion markers. Student’s 2-tailed t-test was used to compare the mean of each group and one way ANOVA was performed with multiple comparison (Bonferroni) testing between the individual timepoints for each group in our cohort. Statistical analysis was performed on data using FlowJo v12, SPSS V25 and R.

#### Tumour biopsies

Duplicate tumour biopsies were collected at baseline, after 2 × cycles of priming (week 9) and after 6 cycles of Avelumab (week 22) if metastatic disease was amenable to biopsy. 4/20 patients had metastatic disease that was amenable to biopsy by palpation or detected by CT at all 3 timepoints. 16/20 patients did not have disease amenable to biopsy at all timepoints. Each of the duplicate biopsies were formalin-fixed, paraffin embedded (FFPE) for HLA-A expression analysis and fresh frozen for RNA-seq and reduced representation bisulfite sequencing (RRBS).

#### HLA-A expression

FFPE biopsies for 4 participants (001, 006, 008, 010) collected at baseline, week 9 and week 22 were confirmed to contain metastatic melanoma using H&E stained sections by a pathologist (REV), with the exception of 001 Week 9 biopsy which did not contain viable melanoma. FFPE tumour biopsies were sliced into 4 μm sections and processed for 3′,3′-diaminobenzidine (DAB) immunohistochemistry using a Ventana Discovery Ultra (Roche, USA) by the Hunter Cancer Biobank. Sections were labelled for HLA-A using recombinant Anti-HLA-A antibody (EP1395Y) (AbCam, USA). All steps from baking to chromogen addition were performed automatically by the Ventana Discovery Ultra. Tissue sections were baked to slides and deparaffinized, and antigen retrieval then occurred at 95°C/pH 9 with a total incubation time of 24 minutes prior to the addition of the primary antibody. Addition of the primary antibody was followed by a 32-minute incubation at 36°C. Slides were then incubated with secondary antibody Anti-Rabbit HQ (Roche) for 20 minutes at 36°C. Slides were digitally scanned using the Aperio™ Digital AT2 Pathology System (Leica Biosystems, Australia) for analysis.

#### RNA-Seq

Frozen biopsies for 3 participants (006, 008, 010) collected at baseline, week 9 and week 22 were confirmed to contain metastatic melanoma using H&E stained sections by a pathologist (REV), with the exception of patient 010 Week 22 biopsy which did not contain viable melanoma. RNA was extracted from biopsies using the AllPrep DNA/RNA/Protein Mini Kit (Qiagen, USA) as per manufacturer’s instructions. Rin score was determined by High Sensitivity RNA Screen Tape analysis (Agilent, USA) and RNA was quantified using Qubit HS. KAPA RNA HyperPrepKit with RiboErase was used for total RNA library prep with rRNA depletion and the 8 RNA libraries were sequenced on NovaSeq. RNA-seq library prep and NovaSeq was provided by the Garvan Institute of Medical Research Kinghorn Centre for Clinical Genomics Sequencing Laboratory.

The RNA-seq data was aligned to hg38 using the STAR aligner (*27*). To quantify the expression of genes, featureCounts (*28*) was used to extract read counts for genes from GENCODE annotations. Gene expression was normalised across samples using the trimmed mean of M-values method (*29*) For the analysis of transposable element expression, the REdiscoverTE pipeline was used (*22*). Briefly, this pipeline uses Salmon (*30*) to quantify repetitive elements from RepeatMasker (*31*) that do not overlap with annotated genes and aggregates TE expression at the family and subfamily level.

#### Whole genome bisulfite sequencing (WGBS)

As described for RNA-Seq, frozen biopsies for 3 participants (006, 008, 010) collected at baseline, week 9 and week 22 were confirmed to contain metastatic melanoma, with the exception of patient 010 Week 22 biopsy which did not contain viable melanoma. DNA was extracted from biopsies using the AllPrep DNA/RNA/Protein Mini Kit (Qiagen, USA) as per manufacturer’s instructions. DNA was quantified using Qubit HS (Invitrogen, USA). 8 DNA libraries were bisulfite converted and sequenced on HiSeq by the Garvan Institute of Medical Research Kinghorn Centre for Clinical Genomics Sequencing Laboratory.

The WGBS data was analysed using the Bismark pipeline (*32*) gainst hg38. CpG island annotations were obtained from the UCSC table browser. For gene annotaitons, CpGs were assigned to a gene within 1kb of its transcription start site, and the final methylation value was the averaged. For the methylation of TE, CpGs that overlapped with RepeatMasker TEs were averaged to obtain methylation values for each TE family.

#### Flow cytometry

Immune cell subsets from peripheral blood collection at baseline, after 2 x cycles of azacitidine and carboplatin (week 9) and after 6 cycles of Avelumab (week 22) were determined using Flow Cytometry on the BD Science (Becton, Dickinson and Company Frankline Lakes USA) Fortessa X20 as previously described (*33, 34*). Frozen PBMC’s were thawed and stained with the following targets: horizon Fixable Viability stain 575V, Hu CD3 BUV737 UCHT1, Hu CD4 BUV496 SK3, Hu CD8 APC-H7 SK1, Hu CD279(PD-1) BB515 EH12.1, Hu TIM-3(CD366) Alexa 647 7D3, Hu LAG-3(CD223) APC-R700 T47-530. Samples were gated to acquire 50,000 live cell events and for lymphocytes, single cells and then live cells. The CD3 subset was gated as the fluorophore (BUV737) vs side scatter (SSE). To separate into CD4+ and CD8+ subsets, all positive CD3+ cells were then further divided into CD4+ and CD8+ by gating CD3 vs CD4+ (BUV496) or CD8+ (APC-H7 SK1). For each of the other surface markers (eg TIM3), positive CD4 or CD8 were gated against the other surface markers eg CD4+ (BUV496 SK3) vs TIM-3 (Alexa 647 7D3). Statistical analysis of immune cell subsets was performed using R and SPSS. A p-value of <0.05 was considered statistically significant. For multiple testing correction Benjamini-Hochberg false discovery rate (FDR) was calculated.

## Data Availability

All data produced in the present study are available upon reasonable request to the authors

## Supplementary Material

Supplementary Figure 1. Whole blood cell counts at baseline, week 9 and week 22

Supplementary Figure 2. Transposable element methylation and expression in patient samples.

## Acknowledgments

The authors would like to acknowledge the participants that consented to be included in this study and the clinical trial staff involved in conducting the study at the Calvary Mater Newcastle and Cairns Hospital, Australia.

## Author contributions

Author’s contributions to the paper listed by CRediT role:

Conceptualization: AvdW, NAB

Methodology: AvdW, SR, LP, NAB

Investigation: AvdW, ML, MCG, REV, NAB

Data curation: AvdW, ML, MCG, RL, AM, REV, NAB

Formal Analysis: MCG, XZ, JWHW, REV, NAB

Funding acquisition: AvdW, NAB

Project administration: NAB

Writing – original draft: AvdW, NAB

Writing – review & editing: AvdW, MCG, JWHW, NAB

