## Supplementary figures for "Repurposing azacitidine and carboplatin to prime for anti-PDL1 re-challenge of immunotherapy-resistant melanoma"

**Supplementary Figure 1. Whole blood cell counts at baseline, week 9 and week 22**

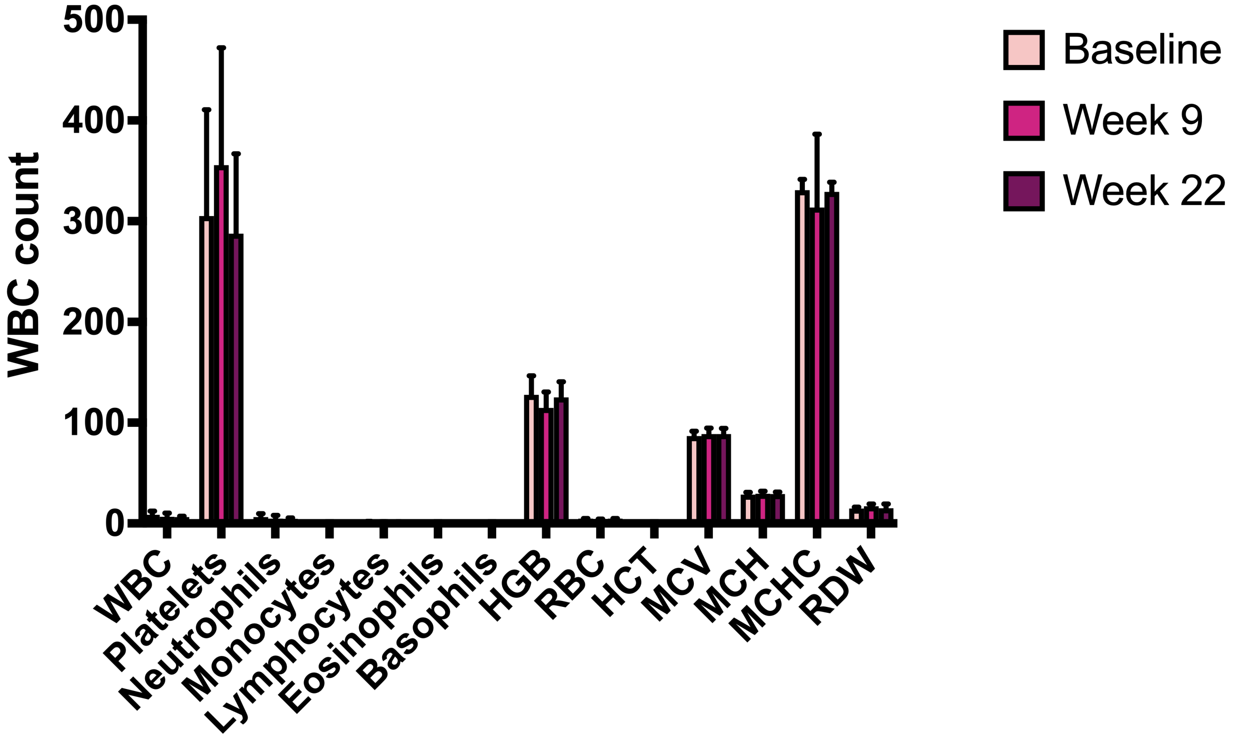

**Supplementary Figure 2. Transposable element methylation and expression in patient samples.**

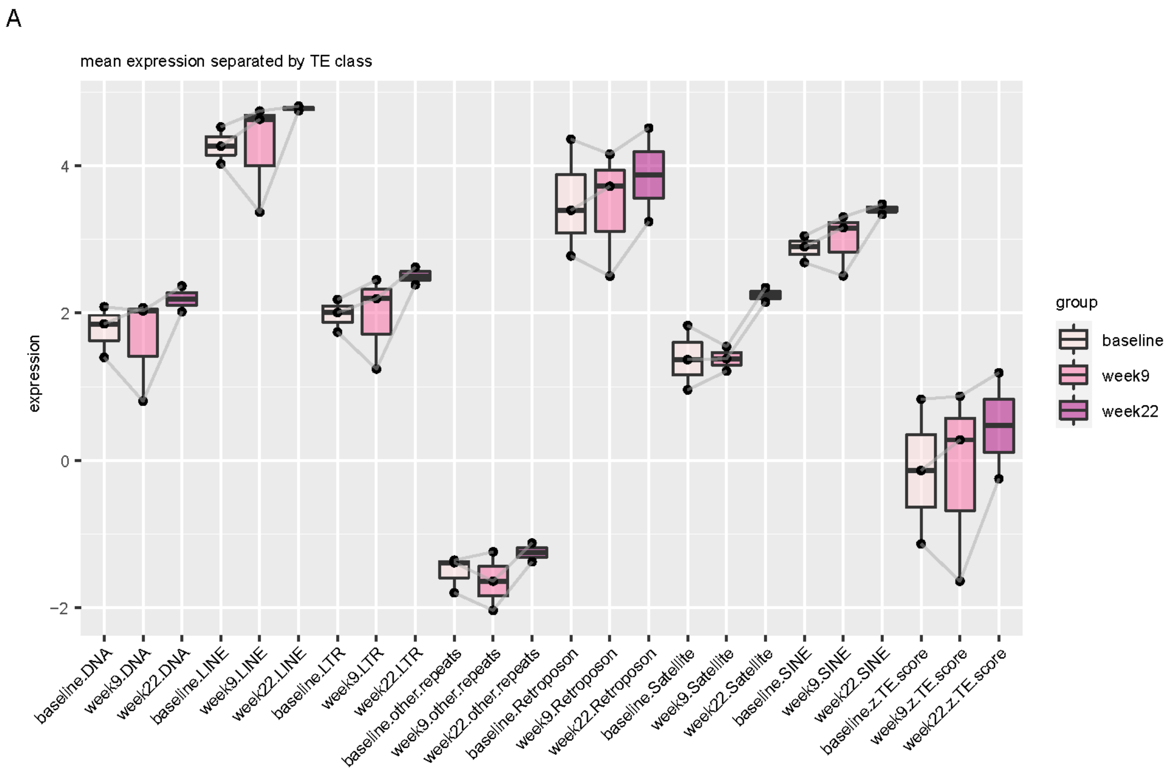


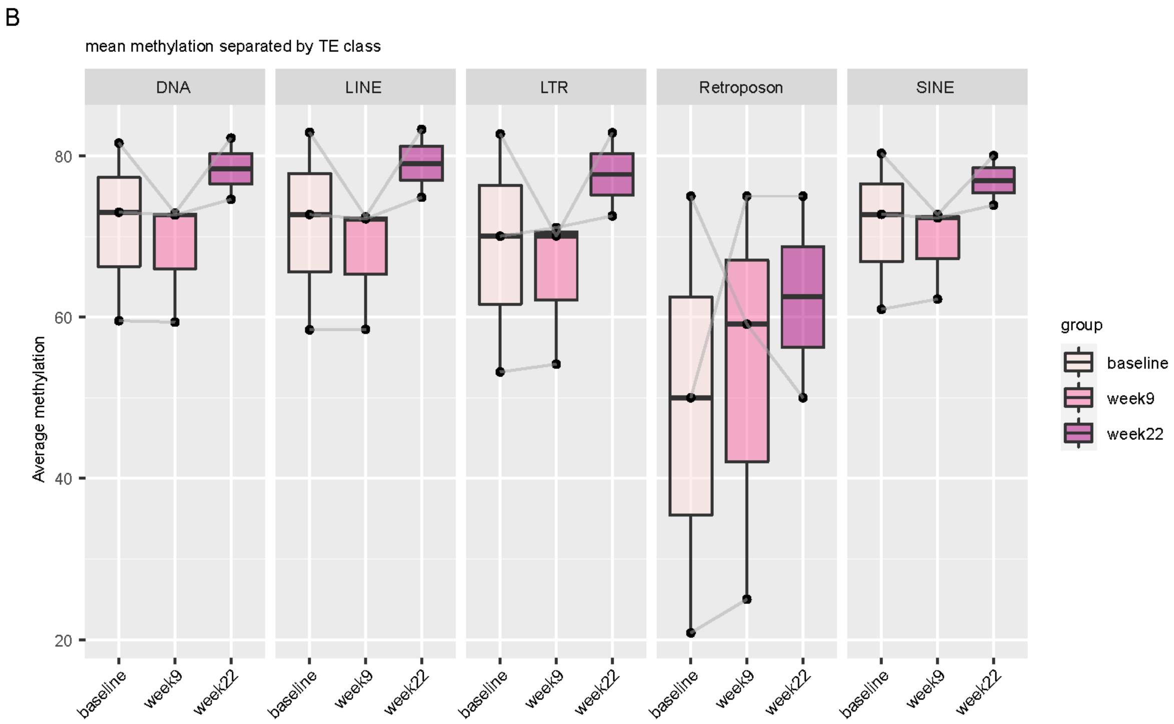
